# Association of poor adherence to guideline-directed medical treatment with adverse events including fatal arrhythmia in heart failure with reduced ejection fraction: A nationwide cohort analysis

**DOI:** 10.64898/2025.12.15.25342327

**Authors:** Moojun Kim, Dong-Hyuk Cho, Jimi Choi, Jong-Chan Youn, Jung-Woo Son, Jooyeon Lee, Taeil Yang, Jaewon Oh, Chan Joo Lee, Byung-Su Yoo, Seok-Min Kang

## Abstract

**Background:** Poor adherence to guideline-directed medical therapy (GDMT) in patients with heart failure (HF) with reduced ejection fraction is associated with higher mortality and hospitalization. However, its association with malignant arrhythmias and aborted sudden cardiac death (SCD) remains unclear. This study assessed the association between GDMT adherence and these outcomes in patients with an implantable cardioverter-defibrillator (ICD) or cardiac resynchronization therapy with a defibrillator (CRT-D).

**Methods:** Patients who had an ICD or CRT-D implanted for primary prevention were included. Those with sustained ventricular tachycardia (VT), ventricular fibrillation/flutter (VF/VFL), or an aborted SCD device before implantation were excluded. Adherence to renin-angiotensin system blockers (RASBs), beta blockers (BBs), and mineralocorticoid receptor antagonists (MRAs) was assessed and categorized as optimal, suboptimal, or poor. The primary outcome was a composite of all-cause mortality, sustained VT, VF/VFL, and aborted SCD.

**Results:** Among 3,780 patients, the prescription rates were 87.5% for RASBs, 89.7% for BBs, and 74.2% for MRAs. Compared with optimal adherence, both suboptimal and poor adherence were associated with an increased risk of the primary outcome. The adverse effects of poor adherence were most evident in patients with ischemic HF. Although lower adherence was correlated with more arrhythmic events, these associations were inconsistent when clinical factors were considered.

**Conclusions:** Lower GDMT adherence was independently associated with increased mortality despite device therapy, particularly in patients with ischemic HF. Although its relationship with arrhythmic outcomes was unclear, sustained adherence remains critical, underscoring the need for targeted interventions to enhance therapeutic continuity.

## Introduction

Heart failure with reduced ejection fraction (HFrEF) is a major clinical phenotype of heart failure (HF), defined as impaired left ventricular (LV) systolic function, with an ejection fraction below 40%.**^(1–3)^** Despite substantial therapeutic advances in recent decades, patients with HFrEF face considerable risks of adverse outcomes, with sudden cardiac death (SCD) remaining a leading cause of mortality. The underlying mechanisms of SCD in this population are multifactorial and involve progressive LV remodeling, autonomic dysregulation, and a predisposition to malignant ventricular arrhythmias (VAs) such as sustained ventricular tachycardia (VT) and ventricular fibrillation or flutter (VF/VFL).**^(4–7)^** Accordingly, reducing arrhythmic risk is an important objective in patients with HFrEF, with implantable cardioverter-defibrillators (ICDs) and cardiac resynchronization therapy with defibrillators (CRT-Ds) serving as key strategies to reduce mortality from fatal arrhythmic events.**^(6)^**

Guideline-directed medical therapy (GDMT) remains the cornerstone of HFrEF management, with robust evidence demonstrating a reduction in mortality, prevention of HF-related hospitalizations, and mitigation of disease progression.**^(8–11)^** Current guidelines recommend a four-pillar regimen comprising renin-angiotensin system blockers (RASBs)—including angiotensin-converting enzyme inhibitors (ACEis), angiotensin receptor blockers (ARBs), or angiotensin receptor-neprilysin inhibitors (ARNIs)—as well as beta blockers (BBs), mineralocorticoid receptor antagonists (MRAs), and sodium-glucose cotransporter 2 (SGLT2) inhibitors. Beyond hemodynamic improvement, these agents exert anti-arrhythmic effects. RASBs modify LV remodeling and fibrosis, BBs reduce sympathetic tone and myocardial excitability, and MRAs preserve electrolyte balance and further suppress fibrosis.**^(2, 7, 12)^** Nevertheless, GDMT adherence in real-world clinical practice remains suboptimal owing to comorbidities, polypharmacy, clinical inertia, and socioeconomic barriers.**^(13–17)^** Poor adherence is consistently associated with an increased risk of all-cause mortality and HF-related hospitalization.**^(18)^**

Although the prognostic benefits of GDMT adherence in reducing mortality and morbidity are well-established, its relationship with arrhythmic outcomes remains unclear. Given that sustained VT, VF/VFL, and aborted SCD are common terminal events in patients with HFrEF, elucidating the role of GDMT adherence in preventing these events has important clinical implications. Accordingly, in this study, we aimed to evaluate the association between GDMT adherence and the incidence of all-cause mortality, sustained VT, VF/VFL, and aborted SCD in a real-world cohort of patients with HFrEF who underwent ICD or CRT-D implantation for primary prevention.

## Methods

### Data sources

This study used data from the Korean National Health Insurance Service (NHIS) database, a nationwide administrative claims registry that provides universal health coverage for nearly the entire South Korean population. The NHIS database offers detailed patient-level information, including demographic profiles, healthcare utilization, diagnoses, prescription records, and results of routine health screening examinations. **^(19)^** Diagnoses are coded according to the Korean Standard Classification of Diseases (KCD), which is aligned with the International Classification of Diseases, 10th Revision (ICD-10). Mortality data were obtained from the qualification records within the NHIS cohort, as compiled by Statistics Korea. The study protocol was approved by the Institutional Review Board for Human Research of Yonsei University Wonju Severance Christian Hospital (approval number: CR321358). The requirement for informed consent was waived owing to its retrospective design and the use of de-identified data provided by the NHIS (database number: NHIS-2022-1-376). All analyses were conducted in accordance with the ethical principles of the Declaration of Helsinki.

### Study population

We analyzed the NHIS claims data from January 2008 to December 2020 and identified 7,123 patients with HFrEF who underwent ICD or CRT-D implantation for primary prevention. Eligible patients were identified based on HF diagnoses recorded using the ICD-10 codes I50, I42, I11.0, I13.0, I13.1, I13.2, and I25.5. Device implantation was confirmed using the relevant procedural codes for ICD and CRT-D.

From the initial cohort, 3,343 patients were excluded for the following reasons: (1) prior diagnosis of sustained VT (I47.2), VF/VFL (I49.0), or aborted SCD (I46.0, I46.9) before device implantation; (2) age <18 years at the time of device implantation; (3) history of left ventricular assist device implantation or heart transplantation before device implantation; (4) absence of any prescription of RASBs, BBs, or MRAs; and (5) follow-up duration of < 3 months. The final study cohort included 3,780 patients who were prescribed at least one class of GDMT **(Figure 1)**.

**Figure 1.**
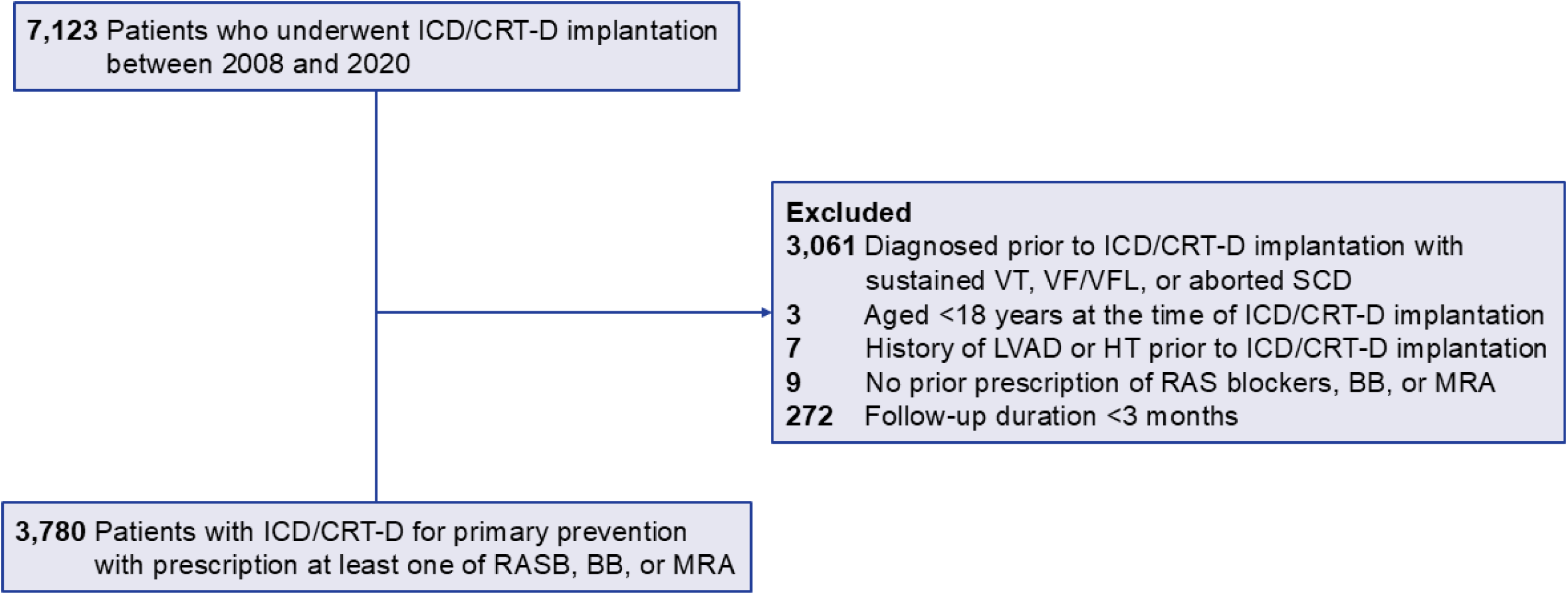
Flow diagram of the study participants. ICD, implantable cardioverter-defibrillator; CRT-D, cardiac resynchronization therapy with defibrillator; VT, ventricular tachycardia; VF, ventricular fibrillation; VFL, ventricular flutter; SCD, sudden cardiac death; RAS, renin-angiotensin system; BB, beta-blocker; MRA, mineralocorticoid receptor antagonist.

The etiology of HF was dichotomized as ischemic or non-ischemic. Ischemic HF was defined as a diagnosis of acute myocardial infarction (I21 and subcodes) or post-myocardial infarction complications (I23 and subcodes) within 1 month before or after HF diagnosis, accompanied by a record of coronary stent insertion. Non-ischemic HF was defined as the absence of these criteria. Patients diagnosed with hypertrophic cardiomyopathy (I42.1, I42.2) were excluded from the subgroup analyses in the non-ischemic HF cohort.

The following comorbidities were identified using KCD codes: hypertension (I10-13 and I15), diabetes mellitus (E11-E14), dyslipidemia (E78), ischemic heart disease (I21, I22, I25.2), atrial fibrillation (I48), cerebrovascular disease (I60-64), peripheral artery disease (I70 and I73) and chronic kidney disease (N18). Socioeconomic status was categorized into low, middle, and high groups based on the health insurance contribution percentiles. Overall comorbidity burden was assessed using the Charlson comorbidity index (CCI).**^(20)^**

### Assessment of GDMT adherence

Adherence to GDMT was assessed using prescription claims data. GDMT was defined as the use of at least one of the following three core pharmacological classes recommended for HFrEF: (1) RASBs, including ACEis, ARBs, or ARNIs; (2) BBs; and (3) MRAs. For each class, adherence was quantified using the proportion of days covered (PDC), which was calculated as the number of days with available prescriptions divided by the total number of follow-up days. As most patients were prescribed more than one class, a weighted PDC approach was used to provide an overall adherence metric. Specifically, we calculated the treatment duration (in days) for each drug class. The class-specific PDC was then weighted by the proportion of the total treatment days attributable to the class, and the weighted values were summed across all three classes to generate a single overall PDC for each patient. Patients were subsequently categorized into three adherence groups: optimal (PDC ≥0.8), suboptimal (0.6 ≤PDC <0.8), and poor (PDC <0.6).

### Study outcomes

The primary outcome was a composite of all-cause mortality, sustained VT, VF/VFL, and aborted SCD during follow-up. Secondary outcomes included each component of the primary outcome and a composite arrhythmic endpoint comprising sustained VT, VF/VFL, and aborted SCD. All-cause mortality was identified using linked death registry data from Statistics Korea, which provides complete nationwide records of death dates and causes. The index date was defined as the date of device implantation.

### Statistical analysis

Categorical variables were presented as absolute numbers with percentages and compared using the chi-square or Fisher’s exact test, as appropriate. Continuous variables with non-normal distributions were reported as medians with interquartile ranges (IQRs) and compared using the Mann–Whitney U test. Comparisons among PDC categories were performed using the Kruskal–Wallis test, followed by pairwise Wilcoxon post-hoc analyses.

A multivariable Cox proportional hazards regression model was used to estimate hazard ratios (HRs) and 95% confidence intervals (CIs) for study outcomes across the PDC categories, adjusting for age, sex, socioeconomic status, etiology of HF comorbidities, and concomitant medications. Incidence rates (IRs) were calculated as the number of events per 100 person-years. Additionally, restricted cubic spline (RCS) Cox regression was used to evaluate potential non-linear associations between continuous PDC for GDMT and study outcomes. A two-sided *P*-value <0.05 was considered statistically significant. All statistical analyses were performed using R software (R Foundation for Statistical Computing, Vienna, Austria).

## Results

### Baseline characteristics

The baseline demographic and clinical characteristics of the patients are presented in **Table 1**. The mean age of the study population was 65.3 ± 12.0 years, and 66.6% were men. Of the 3,780 patients, 1,524 (40.3%) had optimal adherence, 1,169 (30.9%) had suboptimal adherence, and 1,087 (28.8%) had poor adherence. Patients with poor adherence were significantly older than those with optimal adherence, whereas the socioeconomic status was comparable across the adherence groups. Ischemic heart disease was the underlying etiology in 33.8% of patients. Although the CCI was higher in the poor adherence group, other comorbidities were broadly comparable between the groups.

**Table 1.** Baseline characteristics of the study population by guideline-directed medical therapy adherence level.

|  | **Total**<br>**N=3780** | **Optimal**<br>**(PDC≥0.8)**<br>**N=1524** | **Suboptimal**<br>**(0.6≤PDC<0.8)**<br>**N=1169** | **Poor**<br>**(PDC<0.6)**<br>**N=1087** | ***P*** |
| --- | --- | --- | --- | --- | --- |
| **Age** | 65.3 ± 12.0 | 63.8 ± 11.7 | 66.1 ± 11.4 | 66.4 ± 12.8 | <.001 |
| **Women** | 1262 (33.4) | 489 (32.1) | 398 (34.1) | 375 (34.5) | 0.369 |
| **Socioeconomic status** |  |  |  |  | 0.330 |
| Low | 1069 (28.3) |  | 348 (29.8) | 296 (27.2) |  |
| Middle | 1176 (31.1) | 482 (31.6) | 369 (31.6) | 325 (29.9) |  |
| High | 1535 (40.6) | 617 (40.5) | 452 (38.7) | 466 (42.9) |  |
| **Region** |  |  |  |  | 0.143 |
| Metropolitan | 854 (22.6) | 334 (21.9) | 265 (22.7) | 255 (23.5) |  |
| City | 850 (22.5) | 375 (24.6) | 250 (21.4) | 225 (20.7) |  |
| Rural | 2076 (54.9) | 815 (53.5) | 654 (56) | 607 (55.8) |  |
| **Etiology of heart failure** |  |  |  |  | 0.059 |
| Non-ischemic heart disease | 2503 (66.2) | 1017 (66.7) | 744 (63.6) | 742 (68.3) |  |
| Ischemic heart disease | 1277 (33.8) | 507 (33.3) | 425 (36.4) | 345 (31.7) |  |
| **Comorbidities** |  |  |  |  |  |
| Diabetes mellitus | 1120 (29.6) | 437 (28.7) | 378 (32.3) | 305 (28.1) | 0.048 |
| Hypertension | 2627 (69.5) | 1073 (70.4) | 818 (70) | 736 (67.7) | 0.307 |
| Dyslipidemia | 1745 (46.2) | 750 (49.2) | 549 (47) | 446 (41.0) | <.001 |
| Chronic kidney disease | 487 (12.9) | 109 (7.2) | 165 (14.1) | 213 (19.6) | <.001 |
| Atrial fibrillation | 988 (26.1) | 396 (26.0) | 286 (24.5) | 306 (28.2) | 0.136 |
| Left bundle branch block | 358 (9.5) | 161 (10.6) | 110 (9.4) | 87 (8.0) | 0.088 |
| Ischemic heart disease | 2335 (61.8) | 927 (60.8) | 755 (64.6) | 653 (60.1) | 0.054 |
| Cerebrovascular disease | 390 (10.3) | 137 (9.0) | 126 (10.8) | 127 (11.7) | 0.068 |
| Peripheral artery disease | 74 (2.0) | 27 (1.8) | 22 (1.9) | 25 (2.3) | 0.615 |
| Charlson comorbidity index | 4 (2-6) | 3 (2-5) | 4 (3-6) | 4 (3-6) | <.001 |
| **Medication** |  |  |  |  |  |
| Renin-angiotensin system blocker | 3306 (87.5) | 1348 (88.5) | 1058 (90.5) | 900 (82.8) | <.001 |
| Prescription days, median (IQR) | 309 (164-356) | 310 (169.5-358) | 317 (177-357) | 299.5 (142-353) | – |
| Angiotensin receptor-neprilysin inhibitor | 948 (25.1) | 553 (36.3) | 278 (23.8) | 117 (10.8) | <.001 |
| Prescription days, median (IQR) | 185 (84-334.5) | 189 (84-339) | 168 (96-317) | 198 (58-326) | – |
| Aldosterone antagonist | 2803 (74.2) | 1381 (90.6) | 785 (67.2) | 637 (58.6) | <.001 |
| Prescription days, median (IQR) | 263 (115-354) | 310 (153-360) | 228 (103-345) | 184 (66-337) | – |
| Beta blocker | 3390 (89.7) | 1462 (95.9) | 1054 (90.2) | 874 (80.4) | <.001 |
| Prescription days, median (IQR) | 328.5 (181-360) | 340.5 (209-363) | 333 (194-360) | 287 (118-353) | – |
| Calcium channel blocker | 585 (15.5) | 198 (13.0) | 177 (15.1) | 210 (19.3) | <.001 |
| Diuretics except aldosterone antagonist | 3187 (84.3) | 1324 (86.9) | 989 (84.6) | 874 (80.4) | <.001 |
| Digoxin | 792 (21.0) | 309 (20.3) | 246 (21.0) | 237 (21.8) | 0.637 |
| Statin | 2249 (59.5) | 919 (60.3) | 727 (62.2) | 603 (55.5) | 0.004 |

Data are presented as numbers (%) or medians (IQR) unless otherwise specified.
RAS, renin-angiotensin system; ARNI, angiotensin receptor-neprilysin inhibitor; PDC, proportion of days covered.
*P* value resulted from the ANOVA test comparing three groups.

### Medication adherence and prescription patterns

Over a median follow-up period of 3.2 years (IQR, 1.7–5.2 years), 87.5% of the patients received RASBs, with a median prescription duration of 309 days (IQR, 164–356 days). BB was the most frequently prescribed GDMT class (89.7%; median, 328.5 days [IQR, 181–360 days]), followed by MRA (74.2%; median, 263 days [IQR, 115–354 days]). Despite the high overall use of RASBs, the widespread prescription of ARNIs remains limited (25.1%), reflecting their subsequent incorporation into clinical guidelines and gradual adoption in practice.

The patients with optimal adherence had significantly longer prescription durations across all GDMT classes. Regarding prescription rates, the use of RASBs was similar across the adherence groups, whereas BBs and MRAs were more frequently prescribed in patients with optimal adherence than in those with lower adherence **(Table 1)**.

### Clinical outcomes based on GDMT adherence

**Table 2** summarizes the IR and adjusted HR for clinical outcomes stratified by the GDMT adherence level. The IR of the primary composite outcome was 5.91 per 1,000 person-years in the optimal adherence group, increasing to 7.95 and 10.16 per 1,000 person-years in the suboptimal and poor adherence groups, respectively. After adjustment for demographic factors, HF etiology, comorbidities, and concomitant medications, both suboptimal adherence (adjusted HR, 1.24; 95% CI: 1.05–1.45; *P* = 0.009) and poor adherence (adjusted HR, 1.54; 95% CI: 1.31–1.82; *P* <0.001) were independently associated with increased risk compared to optimal adherence.

**Table 2.** Incidence rates and hazard ratios for study outcomes by guideline-directed medical therapy adherence level.

|  | **Optimal**<br>**(PDC≥0.8)**<br>**N=1524** | **Suboptimal**<br>**(0.6≤PDC<0.8)**<br>**N=1169** | ***P*** | **Poor**<br>**(PDC<0.6)**<br>**N=1087** | ***P*** |
| --- | --- | --- | --- | --- | --- |
| **Primary outcome** |  |  |  |  |  |
| Incidence rate | 293 (5.91) | 333 (7.95) |  | 425 (10.16) |  |
| Crude HR (95% CI) | 1 (Ref.) | **1.34 (1.15-1.57)** | **<.001** | **1.72 (1.48-2.00)** | **<.001** |
| Adjusted HR (95% CI) | 1 (Ref.) | **1.24 (1.05-1.45)** | **0.009** | **1.54 (1.31-1.82)** | **<.001** |
| **All-cause mortality** |  |  |  |  |  |
| Incidence rate | 220 (4.21) | 285 (6.44) |  | 379 (8.62) |  |
| Crude HR (95% CI) | 1 (Ref.) | **1.52 (1.28-1.81)** | **<.001** | **2.03 (1.72-2.40)** | **<.001** |
| Adjusted HR (95% CI) | 1 (Ref.) | **1.41 (1.18-1.69)** | **<.001** | **1.83 (1.52-2.20)** | **<.001** |
| **Sustained VT** |  |  |  |  |  |
| Incidence rate | 97 (1.96) | 84 (2.00) |  | 71 (1.68) |  |
| Crude HR (95% CI) | 1 (Ref.) | 1.01 (0.75-1.35) | 0.958 | 0.82 (0.60-1.12) | 0.213 |
| Adjusted HR (95% CI) | 1 (Ref.) | 1.02 (0.75-1.37) | 0.917 | 0.84 (0.60-1.16) | 0.291 |
| **Ventricular fibrillation/flutter** |  |  |  |  |  |
| Incidence rate | 12 (0.23) | 19 (0.43) |  | 22 (0.51) |  |
| Crude HR (95% CI) | 1 (Ref.) | 1.82 (0.88-3.79) | 0.107 | 2.03 (0.99-4.18) | 0.054 |
| Adjusted HR (95% CI) | 1 (Ref.) | 1.66 (0.79-3.51) | 0.184 | 1.81 (0.90-3.65) | 0.096 |
| **Aborted sudden cardiac death** |  |  |  |  |  |
| Incidence rate | 45 (0.86) | 63 (1.43) |  | 54 (1.24) |  |
| Crude HR (95% CI) | 1 (Ref.) | **1.60 (1.09-2.34)** | **0.016** | 1.29 (0.86-1.92) | 0.212 |
| Adjusted HR (95% CI) | 1 (Ref.) | 1.46 (0.99-2.17) | 0.058 | 1.07 (0.68-1.68) | 0.771 |
| **VT + VF + Aborted SCD** |  |  |  |  |  |
| Incidence rate | 146 (2.96) | 159 (3.82) |  | 153 (3.69) |  |
| Crude HR (95% CI) | 1 (Ref.) | **1.28 (1.02-1.60)** | **0.030** | 1.19 (0.94-1.49) | 0.144 |
| Adjusted HR (95% CI) | 1 (Ref.) | 1.24 (0.99-1.57) | 0.063 | 1.12 (0.87-1.44) | 0.383 |

Adjusted for age, sex, socioeconomic status, heart failure etiology, comorbidities, and concomitant medications.
HR, hazard ratio; CI, confidence interval; VT, ventricular tachycardia; VF, ventricular fibrillation; SCD, sudden cardiac death; PDC, proportion of days covered.

A similar trend was observed for all-cause mortality, with corresponding IRs of 4.21, 6.44, and 8.62 per 1,000 person-years in the optimal, suboptimal, and poor adherence groups, respectively. Similarly, in fully adjusted models, both suboptimal (HR, 1.41; 95% CI: 1.18–1.69; *P* <0.001) and poor adherence (HR, 1.83; 95% CI: 1.52–2.20; *P* <0.001) remained significantly associated with increased mortality risk.

In contrast, no significant association was found between GDMT adherence and the incidence of sustained VT or VF/VFL after adjusting for covariates. Among patients with poor adherence, the incidence of arrhythmic events was comparable to that in the optimal adherence group. Sensitivity analyses that did not account for competing risks yielded broadly similar findings; however, a consistent pattern emerged in which poor adherence was associated with a tendency toward high arrhythmic risk **(Table S1–S3)**.

### RCS analysis of GDMT adherence and clinical outcomes

RCS regression models were used to characterize the continuous relationship between GDMT adherence, as quantified by the weighted PDC, and adverse clinical outcomes. A progressive decline in the adjusted HR for both the primary composite outcome **(Figure 2)** and all-cause mortality **(Figure 3)** was observed with increasing adherence. Notably, the risk reduction became more pronounced beyond a PDC threshold of approximately 80%, suggesting a non-linear association, whereby higher levels of adherence were associated with disproportionately greater clinical benefits.

**Figure 2.**
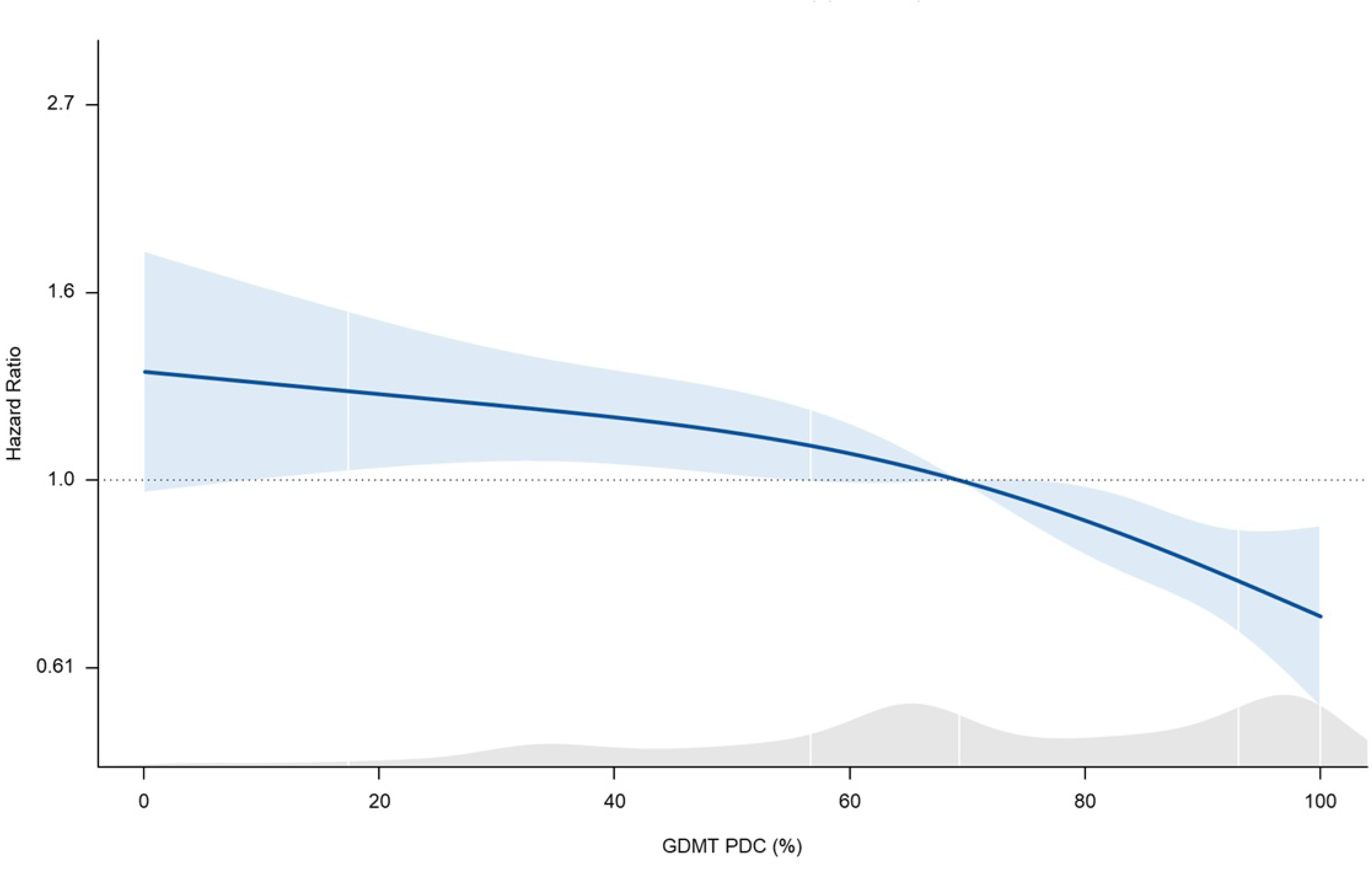
Restricted cubic spline analysis of the association between the proportion of days covered and adjusted hazard ratio of the primary composite outcome. Adjusted hazard ratios and 95% confidence intervals are plotted across the full range of proportion of days covered, using multivariable Cox regression with restricted cubic splines. The reference value was set at PDC = 1.0. GDMT, guideline-directed medical therapy; PDC, proportion of days covered.

**Figure 3.**
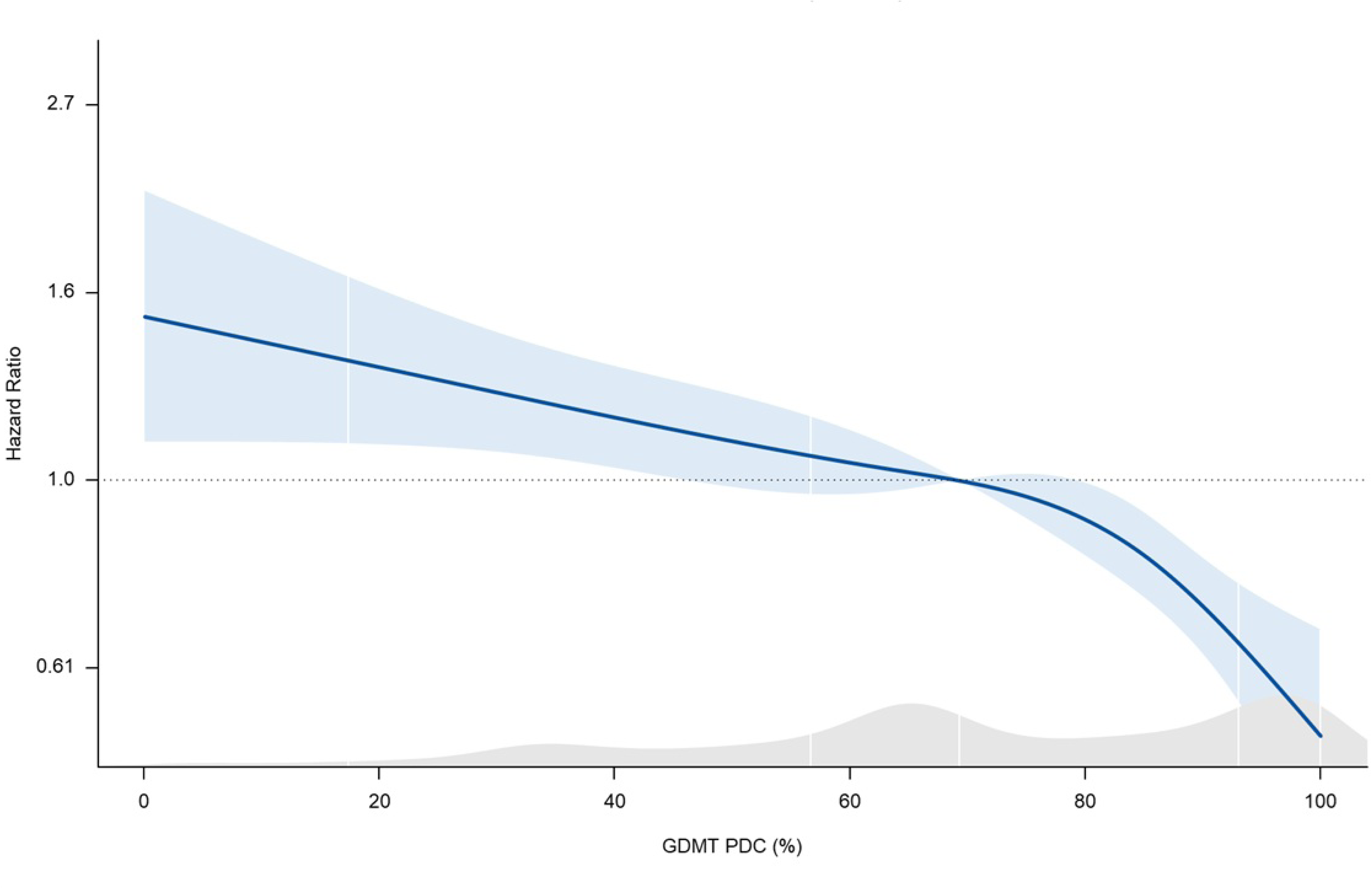
Restricted cubic spline analysis of the association between the proportion of days covered and the adjusted hazard ratio of the all-cause mortality. Adjusted hazard ratios and 95% confidence intervals are plotted across the full range of proportion of days covered, using multivariable Cox regression with restricted cubic splines. The reference value was set at PDC = 1.0. GDMT, guideline-directed medical therapy; PDC, proportion of days covered.

### Subgroup analysis based on baseline characteristics

Subgroup analyses were conducted to assess potential effect modifications across key clinical variables, including HF etiology, age, sex, comorbidity burden, diabetes, and hypertension.

First, the association between GDMT adherence and adverse outcomes differed according to the HF etiology **(Table 3)**. In patients with ischemic HF, both suboptimal and poor adherence were associated with a markedly increased risk of the primary composite outcome and all-cause mortality. Poor adherence was associated with an adjusted HR of 1.95 (95% CI: 1.49–2.55) for the primary composite outcome and 2.26 (95% CI: 1.67–3.05) for all-cause mortality. These associations were significantly stronger than those observed in patients with non-ischemic HF, with significant interaction terms for both outcomes (*P* for interaction = 0.002 and 0.009, respectively).

**Table 3.** Subgroup analysis of study outcomes by heart failure etiology.

|  | Non-ischemic |  |  |  |  | Ischemic |  |  |  |  | *P* value for difference* |
| --- | --- | --- | --- | --- | --- | --- | --- | --- | --- | --- | --- |
|  | Total / Event (IR) | Crude HR (95% CI) | *P* | Adjusted HR (95% CI) | *P* | Total / Event (IR) | Crude HR (95% CI) | *P* | Adjusted HR (95% CI) | *P* |  |
| **Primary outcome** |  |  |  |  |  |  |  |  |  |  |  |
| Optimal | 1017 / 197 (5.82) | 1 (Ref.) |  | 1 (Ref.) |  | 507 / 96 (6.12) | 1 (Ref.) |  | 1 (Ref.) |  |  |
| Suboptimal | 744 / 197 (7.22) | **1.24 (1.02-1.51)** | **0.033** | 1.15 (0.94-1.41) | 0.171 | 425 / 136 (9.30) | **1.51 (1.18-1.95)** | **0.001** | **1.42 (1.10-1.84)** | **0.008** | 0.094 |
| Poor | 742 / 258 (8.42) | **1.44 (1.20-1.74)** | **<.001** | **1.36 (1.12-1.66)** | **0.002** | 345 / 167 (14.93) | **2.44 (1.90-3.14)** | **<.001** | **1.95 (1.49-2.55)** | **<.001** | **0.002** |
| **All-cause mortality** |  |  |  |  |  |  |  |  |  |  |  |
| Optimal | 1017 / 148 (4.13) | 1 (Ref.) |  | 1 (Ref.) |  | 507 / 72 (4.38) | 1 (Ref.) |  | 1 (Ref.) |  |  |
| Suboptimal | 744 / 163 (5.64) | **1.36 (1.09-1.69)** | **0.007** | **1.27 (1.01-1.59)** | **0.040** | 425 / 122 (7.93) | **1.79 (1.35-2.38)** | **<.001** | **1.70 (1.28-2.27)** | **<.001** | **0.020** |
| Poor | 742 / 224 (6.96) | **1.66 (1.35-2.05)** | **<.001** | **1.63 (1.31-2.03)** | **<.001** | 345 / 155 (13.17) | **2.99 (2.25-3.96)** | **<.001** | **2.26 (1.67-3.05)** | **<.001** | **0.009** |
| **Sustained VT** |  |  |  |  |  |  |  |  |  |  |  |
| Optimal | 1017 / 68 (2.00) | 1 (Ref.) |  | 1 (Ref.) |  | 507 / 29 (1.86) | 1 (Ref.) |  | 1 (Ref.) |  |  |
| Suboptimal | 744 / 51 (1.85) | 0.92 (0.64-1.33) | 0.669 | 0.93 (0.65-1.34) | 0.693 | 425 / 33 (2.27) | 1.19 (0.72-1.95) | 0.498 | 1.22 (0.73-2.04) | 0.455 | 0.700 |
| Poor | 742 / 47 (1.51) | 0.75 (0.52-1.09) | 0.137 | 0.74 (0.50-1.08) | 0.122 | 345 / 24 (2.14) | 0.99 (0.57-1.71) | 0.968 | 1.10 (0.62-1.96) | 0.736 | 0.178 |
| **Ventricular fibrillation/flutter** |  |  |  |  |  |  |  |  |  |  |  |
| Optimal | 1017 / 9 (0.25) | 1 (Ref.) |  | 1 (Ref.) |  | 507 / 3 (0.18) | 1 (Ref.) |  | 1 (Ref.) |  |  |
| Suboptimal | 744 / 12 (0.42) | 1.62 (0.68-3.88) | 0.275 | 1.53 (0.64-3.65) | 0.334 | 425 / 7 (0.46) | 2.41 (0.62-9.37) | 0.203 | 2.12 (0.52-8.58) | 0.293 | 0.925 |
| Poor | 742 / 13 (0.41) | 1.56 (0.66-3.71) | 0.314 | 1.35 (0.58-3.11) | 0.486 | 345 / 9 (0.78) | 3.51 (0.95-13.02) | 0.060 | 3.39 (0.87-13.19) | 0.078 | 0.225 |
| **Aborted sudden cardiac death** |  |  |  |  |  |  |  |  |  |  |  |
| Optimal | 1017 / 31 (0.87) | 1 (Ref.) |  | 1 (Ref.) |  | 507 / 14 (0.85) | 1 (Ref.) |  | 1 (Ref.) |  |  |
| Suboptimal | 744 / 35 (1.22) | 1.36 (0.84-2.21) | 0.209 | 1.26 (0.77-2.06) | 0.367 | 425 / 28 (1.82) | **2.04 (1.08-3.86)** | **0.029** | 1.90 (0.99-3.67) | 0.054 | 0.262 |
| Poor | 742 / 32 (1.00) | 1.08 (0.66-1.78) | 0.761 | 0.94 (0.56-1.60) | 0.832 | 345 / 22 (1.88) | 1.79 (0.91-3.50) | 0.090 | 1.36 (0.66-2.81) | 0.411 | 0.400 |
| **VT + VF + Aborted SCD** |  |  |  |  |  |  |  |  |  |  |  |
| Optimal | 1017 / 99 (2.93) | 1 (Ref.) |  | 1 (Ref.) |  | 507 / 47 (3.04) | 1 (Ref.) |  | 1 (Ref.) |  |  |
| Suboptimal | 744 / 90 (3.31) | 1.13 (0.85-1.50) | 0.409 | 1.11 (0.83-1.48) | 0.499 | 425 / 69 (4.75) | **1.56 (1.08-2.25)** | **0.018** | **1.53 (1.05-2.24)** | **0.028** | 0.056 |
| Poor | 742 / 97 (3.18) | 1.07 (0.81-1.42) | 0.626 | 1.01 (0.75-1.35) | 0.969 | 345 / 56 (5.07) | 1.45 (0.98-2.15) | 0.063 | 1.38 (0.91-2.10) | 0.132 | 0.080 |

Adjusted for age, sex, socioeconomic status, heart failure etiology, comorbidities, and concomitant medications.
VT, ventricular tachycardia; VF, ventricular fibrillation; SCD, sudden cardiac death; IR, incidence rate; HR hazard ratio; CI, confidence interval.

In contrast, GDMT adherence was not clearly associated with either individual arrhythmic endpoints or their composite, regardless of the HF etiology. Although an increased risk of aborted SCD was observed among patients with ischemic HF and suboptimal adherence in the unadjusted analyses, this association was attenuated after adjusting for confounders and was not evident in either the poor adherence group or the composite arrhythmic outcome. Analyses stratified by age, sex, and comorbidity burden demonstrated consistent patterns **(Table S4–S8)**. The detrimental impact of lower GDMT adherence on clinical outcomes appeared more pronounced among clinically vulnerable subgroups, including older adults (≥65 years), women, and patients with a high comorbidity burden (CCI ≥4), with significant interaction effects noted for both the primary composite outcome and all-cause mortality. Notably, among patients with CCI ≥4 or hypertension, poor adherence was associated with an increased risk of VF/VFL.

### Sensitivity analyses using alternative GDMT adherence

To assess the robustness of the primary findings, sensitivity analyses were performed using alternative definitions of adherence. When GDMT adherence was categorized into tertiles, both the middle (PDC, 0.63–0.87) and lowest (PDC <0.63) tertiles were associated with incrementally higher risks of the primary composite outcome and all-cause mortality, relative to the highest adherence group **(Table S9)**. Consistent results were observed with a binary classification, where non-optimal adherence was significantly associated with an increased risk of both outcomes **(Table S10)**. Furthermore, modelling PDC as a continuous variable revealed a dose-response relationship—each 10% increase in PDC was linked to a reduced risk of the primary composite outcome (HR, 0.93; 95% CI: 0.91–0.96) and all-cause mortality (HR, 0.91; 95% CI: 0.88–0.93) **(Table S11)**. The results for arrhythmic outcomes remained consistent across all sensitivity models and aligned with the findings from the primary analysis.

## Discussion

Several key findings were identified in this nationwide cohort study of patients with HFrEF who underwent ICD or CRT-D implantation for primary prevention. First, lower GDMT adherence, as quantified by the weighted PDC, was independently associated with a higher risk of the primary composite outcome, including all-cause mortality and major VA events. Second, this association was more pronounced among patients with ischemic HF, suggesting etiological heterogeneity in the vulnerability to suboptimal pharmacological treatments. Third, although lower adherence was associated with higher crude rates of arrhythmic events, these associations were attenuated after adjusting for the covariates. Lastly, the detrimental effect of poor adherence was particularly notable among clinically vulnerable subgroups, which include older adults, women, and individuals with high comorbidity burden (CCI ≥4). These results reinforce previous evidence underscoring the central role of optimal GDMT in HFrEF, even in patients selected for ICD or CRT-D.**^(21)^** Randomized controlled trials have consistently shown substantial reductions in mortality and HF-related hospitalization with comprehensive pharmacological therapy; however, real-world adherence remains suboptimal owing to multifactorial barriers.**^(13–17)^** This persistent gap in implementation is particularly concerning in the era of device-based therapy, where pharmacological treatment remains essential for mitigating both arrhythmic and non-arrhythmic risks. Evidence from single**^(22)^** and multicentre cohorts**^(23)^** has shown stepwise reductions in mortality with greater use of concurrent GDMT classes, even among patients already receiving device therapy. Our study extends this literature by using a validated, quantitative adherence metric or weighted PDC, and shows that even moderate reductions in adherence confer excess risk. Furthermore, the RCS analyses suggested non-linearity around the conventional 80% threshold, emphasizing the importance of achieving and sustaining high therapeutic continuity.

Despite robust trial evidence, large-scale registries continue to report marked underuse and under-titration of GDMT in routine clinical practice. In the CHAMP-HF registry, fewer than 1% of patients with HFrEF received triple therapy at target doses despite a few contraindications**^(24)^**. Similarly, the CHECK-HF registry in the Netherlands reported that only one-third of patients were prescribed all three core classes, with substantial inter-institutional variation suggestive of practice-level inertia**^(25)^**. The multinational ASIAN-HF registry further showed that common clinical barriers, such as hypotension or bradycardia, did not fully account for the low uptake of GDMT, indicating broad system-level barriers**^(26)^**. Most recently, the EVOLUTION-HF study highlighted persistent therapeutic inertia during initiation and titration, even within integrated health systems**^(27)^**. Our findings extend prior registry-based observations by demonstrating that GDMT is not only underutilized but that poor adherence among patients selected for ICD or CRT-D implantation is independently associated with an increased risk of all-cause mortality. This highlights the fact that prescriptions alone are insufficient, and sustained implementation and adherence remain critical for improving outcomes.

Building upon this, we found that the adverse impact of suboptimal adherence was more pronounced in patients with ischemic HF than in those with non-ischemic etiologies, consistent with prior studies reporting poorer prognosis in ischemic than in non-ischemic cardiomyopathy.**^(28, 29)^** Ischemic HF is characterized by extensive myocardial scarring, residual ischemia, and conduction abnormalities that create a pro-arrhythmic substrate that increases the risk of progressive pump failure and SCD. Neurohormonal blockade achieved with optimal GDMT helps counteract these mechanisms by limiting adverse remodeling, sympathetic overactivity, and electrical instability.**^(30, 31)^** Therefore, sustained adherence is crucial for preserving hemodynamics and reducing arrhythmic vulnerabilities. Previous studies have shown that VA events occur more frequently in patients with ischemic HF, and ICDs provide a greater survival benefit in this subgroup**^(32, 33)^** Therefore, a stronger association between adherence and arrhythmic events may be anticipated in patients with ischemic HF. However, this was not observed in our study, likely reflecting the overall low number of arrhythmic events and the mitigating effects of the device therapy. Taken together, while patients with ischemic HF remain at particularly high risk, improving adherence and ensuring therapeutic continuity in this subgroup may yield substantial prognostic benefits. Patients with suboptimal adherence showed higher rates of VA events and aborted SCD; however, these associations were no longer significant after adjusting for covariates. This may be owing to improved LV function and reverse remodeling through device therapy, as well as the interruption of malignant rhythms via anti-tachycardia pacing or shock delivery, which together could have reduced the likelihood of clinically apparent events being documented in the NHIS claims data.

Lower GDMT adherence was anticipated to be associated with a higher burden of VA events, supported by evidence that BBs reduce SCD in patients with HFrEF.**^(34–36)^** However, in our study, BB use was lower among patients with suboptimal adherence, but the VA incidence did not differ across the adherence groups. One explanation is the relatively short follow-up period, which may have been insufficient to capture adherence-related differences in arrhythmic outcomes, particularly in this primary prevention cohort where events occur slowly. Extended follow-ups from the SCD-HeFT and DANISH trials, both longer than in our study, demonstrated that reductions in arrhythmic mortality became apparent only after a prolonged period.**^(37, 38)^**

While differences in arrhythmic outcomes were not observed, the adverse effects of poor adherence were more pronounced among older adults, women, and those with a high CCI—groups often underrepresented and considered to have a higher risk of medication-related side effects.**^(25, 26)^** In these vulnerable populations, even modest improvements in adherence may lead to significant reductions in morbidity and mortality rates. Accordingly, targeted strategies such as digital health platforms**^(39)^** and pharmacist-led titration programs**^(11, 40)^** have shown promise in supporting long-term adherence. They may be particularly effective in supporting long-term adherence in these groups.

This study had certain limitations. First, as this was an observational study, we cannot conclude with certainty that the observed associations were causal. Second, adherence was estimated from prescription claims data and may not reflect the actual drug intake or the clinical rationale for dose changes or discontinuation. Third, although the PDC metric is a widely used measure of adherence over time, it does not account for dose intensity or treatment sequencing, both of which may influence treatment effectiveness. Fourth, VA events and aborted SCD were identified using claims-based definitions without formal adjudication, raising the possibility of misclassification. Additionally, the adjudication of cause-specific mortality was not possible owing to the limitations inherent to the claims-based nature of the dataset. Finally, as the study period preceded the incorporation of SGLT2 inhibitors into GDMT for HFrEF, their contribution to adherence could not be assessed.

In conclusion, in this large nationwide cohort of patients with HFrEF treated with ICD or CRT-D for primary prevention, lower GDMT adherence was independently associated with a higher risk of all-cause mortality, with more pronounced effects in patients with an ischemic etiology and in clinically vulnerable subgroups. While arrhythmic outcomes appear to be less influenced by adherence, reflecting device-mediated protection, sustained pharmacological optimization remains essential to improve the overall prognosis.

## Data Availability

The datasets used and/or analyzed during the current study are available from the corresponding author on reasonable request.

## Non-standard Abbreviations and Acronyms

ACEis: angiotensin-converting enzyme inhibitors,
ARBs: angiotensin receptor blockers,
ARNIs: angiotensin receptor–neprilysin inhibitors,
BBs: beta blockers,
CCI: Charlson comorbidity index,
CRT-D: cardiac resynchronization therapy with a defibrillator,
GDMT: guideline-directed medical therapy,
HF: heart failure,
HFrEF: heart failure with reduced ejection fraction,
ICD: implantable cardioverter-defibrillator,
ICD-10: International Classification of Diseases, 10th Revision,
IRs: incidence rates,
KCD: Korean Standard Classification of Diseases,
LV: left ventricular,
MRAs: mineralocorticoid receptor antagonists,
NHIS: Korean National Health Insurance Service,
PDC: proportion of days covered,
RASBs: renin–angiotensin system blockers,
RCS: restricted cubic spline,
SCD: sudden cardiac death,
SGLT2: sodium–glucose cotransporter 2,
VAs: ventricular arrhythmias,
VF/VFL: ventricular fibrillation or flutter,
VT: ventricular tachycardia.

## Acknowledgements

None.

## Sources of Funding

None.

## Disclosures

None declared.

## Notes

### Competing Interest Statement

The authors have declared no competing interest.

### Clinical Trial

None.

### Author Declarations

The study protocol was approved by the Institutional Review Board for Human Research of Yonsei University Wonju Severance Christian Hospital (approval number: CR321358).

